# COVID-Classifier: An automated machine learning model to assist in the diagnosis of COVID-19 infection in chest x-ray images

**DOI:** 10.1101/2020.05.09.20096560

**Authors:** Abolfazl Zargari Khuzani, Morteza Heidari, S. Ali Shariati

## Abstract

Chest-X ray (CXR) radiography can be used as a first-line triage process for non-COVID-19 patients with pneumonia. However, the similarity between features of CXR images of COVID-19 and pneumonia caused by other infections make the differential diagnosis by radiologists challenging. We hypothesized that machine learning-based classifiers can reliably distinguish the CXR images of COVID-19 patients from other forms of pneumonia. We used a dimensionality reduction method to generate a set of optimal features of CXR images to build an efficient machine learning classifier that can distinguish COVID-19 cases from non-COVID-19 cases with high accuracy and sensitivity. By using global features of the whole CXR images, we were able to successfully implement our classifier using a relatively small dataset of CXR images. We propose that our COVID-Classifier can be used in conjunction with other tests for optimal allocation of hospital resources by rapid triage of non-COVID-19 cases.

## Introduction

Chest-X ray (CXR) radiography is one of the most commonly used and accessible methods for rapid examination of the lung conditions [1]. CXR images are almost immediately available for analysis by radiologists. The availability of CXR radiography made it one of the first imaging modalities to be used during the recent COVID-19 pandemic. In addition, the rapid CXR turnaround was used by the radiology departments in Italy and the U.K. to triage non-COVID-19 patients with pneumonia to allocate hospital resources efficiently [2]. However, there are many common features between medical images of COVID-19 and pneumonia caused by other viral infections such as common flu (Influenzas A) [2]. This similarity makes a differential diagnosis of COVID-19 cases by expert radiologists challenging [2, 3]. A reliable automated algorithm for classification of COVID-19 and non-COVID-19 CXR images can speed up the triage process of non-COVID-19 cases and maximize the allocation of hospital resources to COVID-19 cases.

Machine learning (ML) based methods have shown unprecedented success in the reliable analysis of medical images [4-8]. ML-based approaches are scalable, automatable, and easy to implement in clinical settings [9, 10]. A common application of ML-based image analysis is the classification of images with highly similar features. This approach relies on the segmentation of image region of interest, identification of effective image features computed from the segmented area in the spatial or frequency domain, and development of an optimal machine learning-based classification method to accurately assign image samples into target classes [11]. Here, we hypothesized that CXR images of COVID-19 patients can be reliably distinguished from other forms of pneumonia using an ML-based classifier. We used a dimensionality reduction approach to generate a model with an optimized set of synthetic features that can distinguish COVID-19 images with an accuracy of 94% from non-COVID-19 cases. A distinct feature of our model is identification and fusion of the global image features computed from the whole CXR image without lesion segmentation, which enables us to generate a new quantitative imaging marker for predicting the likelihood of a testing case being COVID-19. This new global X-ray image feature-based approach not only avoids lesion segmentation but also reduces the requirement of large training dataset as is the case for the conventional deep learning approach. Our study provides strong proof of concept that simple ML-based classification can be efficiently implemented as an adjunct to other tests to facilitate differential diagnosis of CXR images of COVID-19 patients. More broadly, we think that our approach can be easily implemented in any future viral outbreak for the rapid classification of CXR images.

## Results

### Generation of synthetic features

Identification of optimal features of the CXR images can decrease the feature space of ML models by generating key correlated synthetic features and removing less important features. These synthetic features perform more reliably in classification tasks while reducing the size of the ML models. Importantly, a more robust ML classifier can be generated by increasing the ratio between the training cases per class and image features. We initially extracted 252 features from the whole CXR image without involving lesion segmentation (Fig 1 A and Supplementary Figure 1) to finally generate a feature pool from 420 CXR images (Fig1 B). We hypothesized that we can use a feature analysis scheme to reduce the size of the feature space to an optimal number of features. We computed Pearson correlation coefficients resulting in a matrix for each pairwise feature combination (Fig1 C). Analysis of the histograms of the initial feature pool shows that more than 73% of features have correlation coefficients of less than 0.4 (Fig1 D), indicating that the feature pool created in our study has provided a comprehensive view of the cases, containing relatively small redundancy. We used Kernel-Principal Component Analysis (PCA) method to reduce the dimensionality of the feature space to an optimal number of synthetic features composed of correlated features. By employing PCA, we converted the original pool 252 features to 64 new synthetic features resulting in a ~4x smaller feature space. This vector of 64 selected features was used for classification purposes.

**Figure 1:**
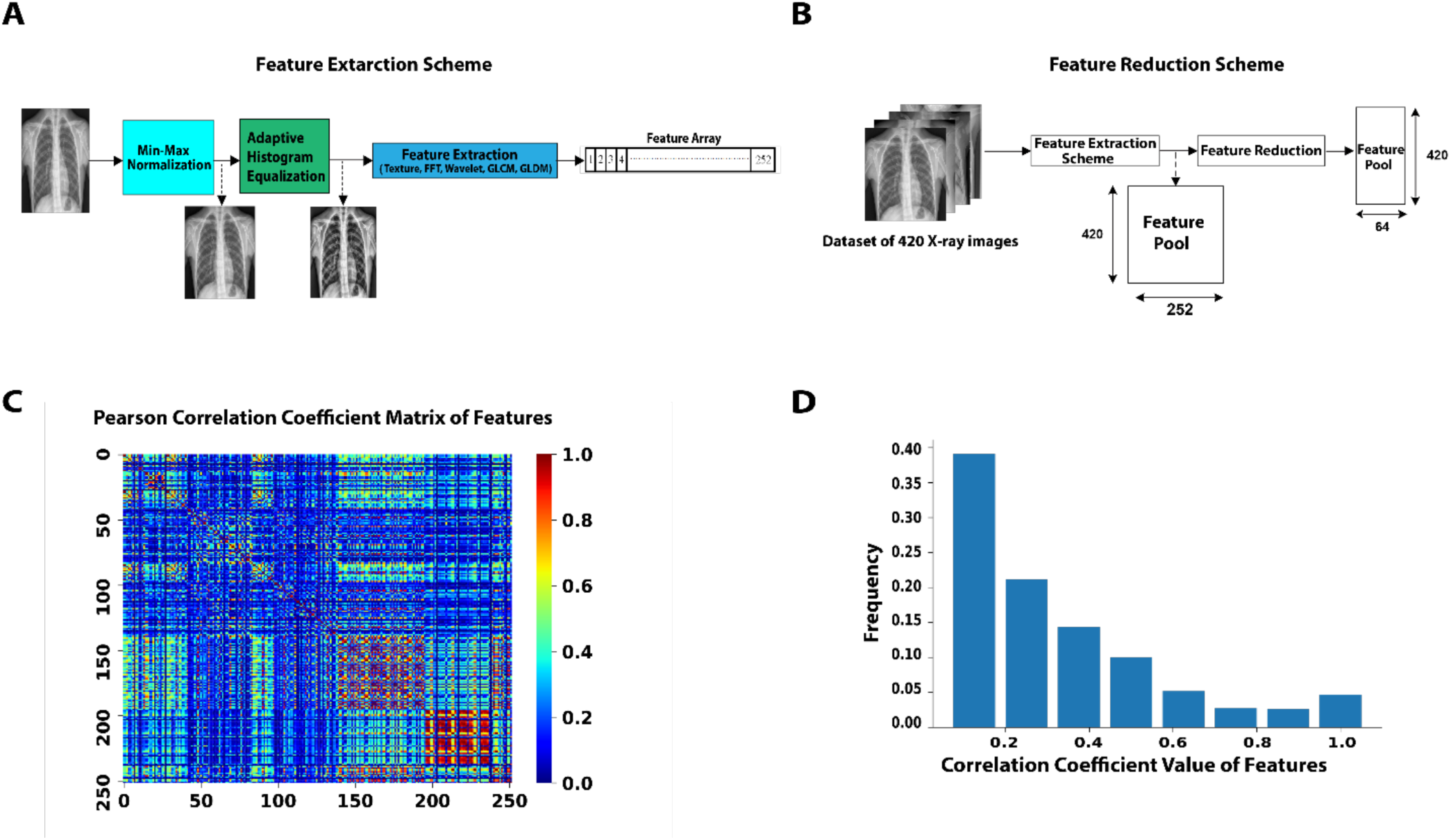
A) Feature extraction scheme to construct a feature array for each CXR image using the Texture, FFT, Wavelet, GLCM, and GLDM methods (See method section for the description of the features). B) A schematic diagram of creating a feature pool for 420 CXR images and applying a feature reduction method. C, D) Correlation analysis of features. The heat map (C) and histogram representation (D) of the Pearson correlation coefficients.

### Classification Performance

To design our classifier, we grouped our CXR images into three target classes, each containing 140 images; normal, COVID-19, non-COVID-19 pneumonia (Supplementary Figure 2). We used a multi-layer neural network with two hidden layers and one output classifier to classify CXR images into three groups (Fig 2). During training our model, both training and validation sets reached ~ 0.22 loss score and 94% accuracy after 33 epochs (Fig 3 A). The loss graph showed a good fit between validation and training curves, confirming that our model is not suffering from overfitting or underfitting. We would like to note that our model has ~10,000 parameters that are considerably smaller than typical images classification models such as AlexNET with 60 million parameters [12], VGG-16 with 138 million [13], GoogleNet-V1 with 5 million [14], and ResNet-50 with 25 million parameters [15]. Next, we generated a receiver operating characteristic (ROC) curve and computed area under the ROC (AUC) to further assess the performance of our model (Fig3 B). A comparison of CXR images of COVID-19 cases with non-COVID-19 showed that our model has100% sensitivity and 96% precision when evaluated on a test set of 84 CXR images (Fig3 C and Table 1). Moreover, our synthetic feature classifier outperforms any single feature classifier as measured by AUC (Fig3 D). It is noteworthy that single synthetic features as the primary fast and low computational cost classifier can be accurate up to ~ 90% (Supplmenatray Figure 3).

**Figure 2:**
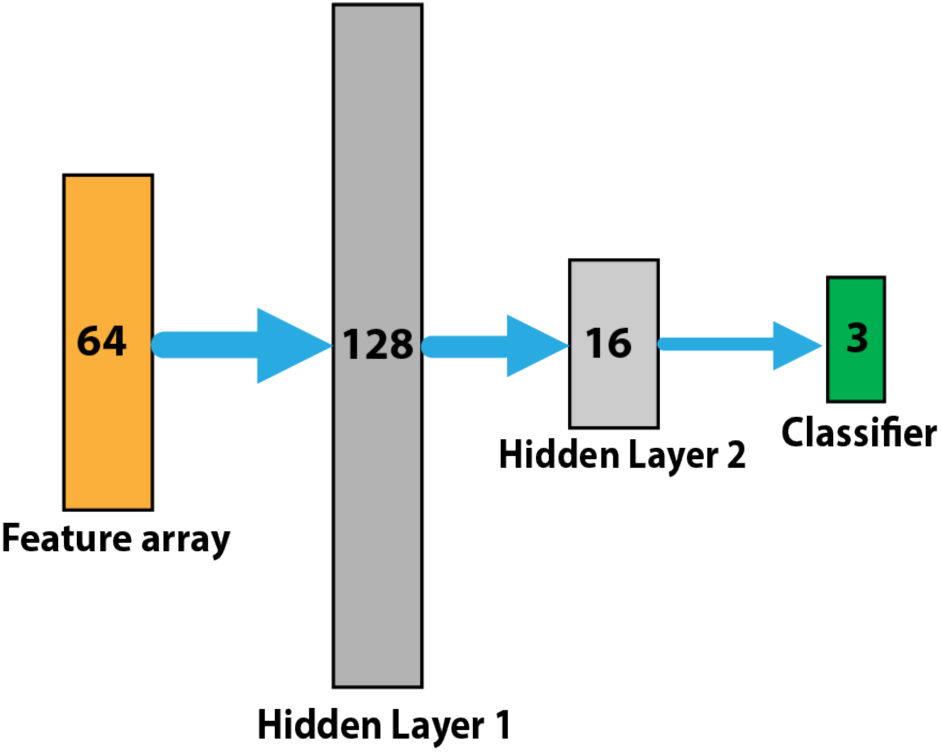
Multi-layer neural network designed for the classification task including two hidden layers with 128 and 16 neurons respectively and a final classifier to classify cases into three categories of normal, COVID-19, non-COVID-19 pneumonia

**Figure 3:**
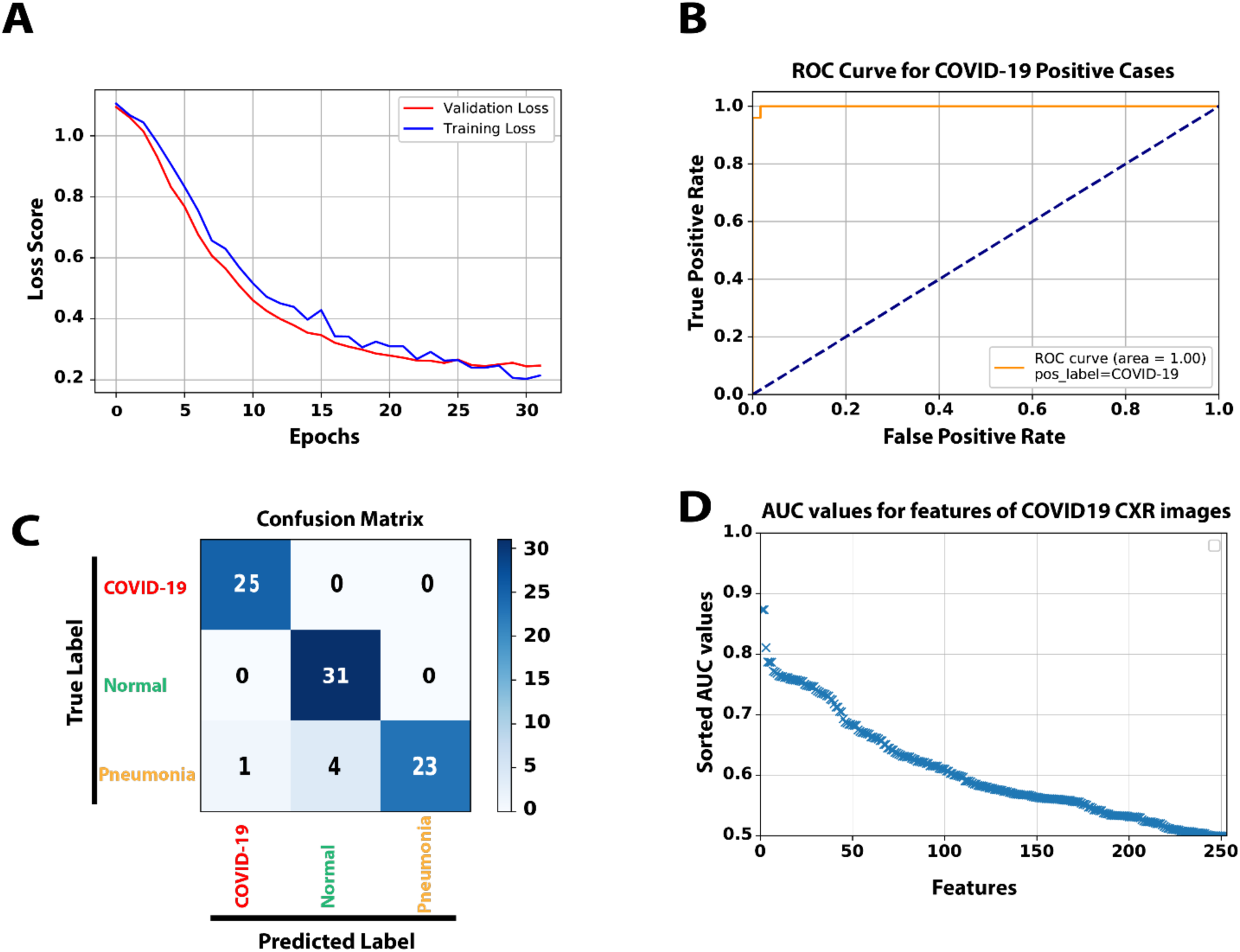
A) The loss score graph of the training and validation sets during the model training process. B) The ROC curve generated from 84 test samples, while COVID-19 is the target class. C) The Confusion matrix of predicting 84 test samples in three categories. D) To compares and analyze the discrimination power of different single features among the original 252 extracted features, we used AUC values as an indicator. All features were sorted in the order of their AUC values.

**Table 1:**
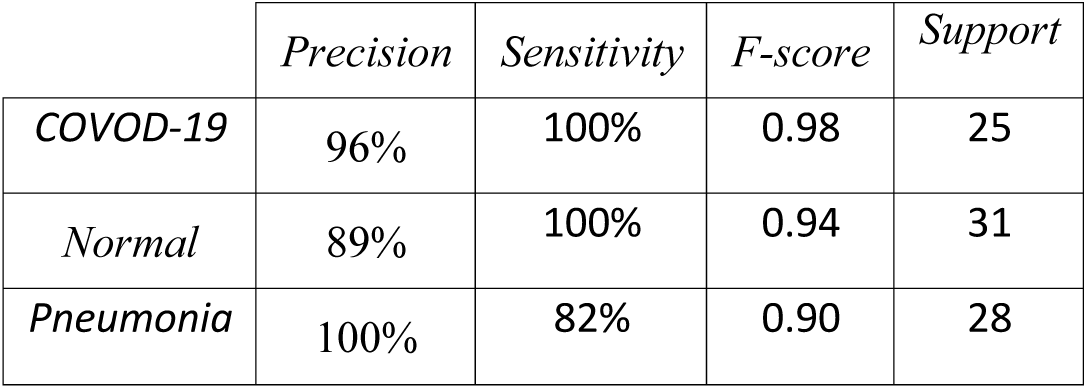
Assessment of evaluation metrics for three target class labels using 84 test samples

|  | *Precision* | *Sensitivity* | *F-score* | *Support* |
| --- | --- | --- | --- | --- |
| *COVID-19* | 96% | 100% | 0.98 | 25 |
| *Normal* | 89% | 100% | 0.94 | 31 |
| *Pneumonia* | 100% | 82% | 0.90 | 28 |

## Discussion

In this study, we demonstrated that efficient machine learning classifier can accurately distinguish COVID-19 CXR images from normal cases and also pneumonia caused by other viruses. Although different imaging modalities have been applied for lung screening [16-18], X-ray remains the fastest and widely used tool for population-based lung disease screening. However, a large number of suspicious lung lesions can result in misclassification of cases. Thus, the development of new approaches to facilitate the classification of different types of lung conditions is crucial to improve the efficacy of lung screening and analysis. In this study, we developed a novel machine learning scheme utilizing the global image features to predict the probability of the testing cases being COVID-19 without lesion segmentation. Our work has a number of new observations as follows:

First, instead of computing image features from the segmented area, we extracted the global image features from the whole chest area, which avoids the difficulty and errors in lesion segmentation and finding the optimal size of the ROIs to include the lesions with varying sizes and shapes. Our result indicates that the clinically meaningful information is not only focused on the lesion but also distributes on the entire chest area of the X-ray image.

Second, unlike many previously developed machine learning models that focus on computing the texture-based features in the spatial domain, we calculated image features in both the spatial domain (Texture, GLDM, GLCM) and frequency domain (FFT and Wavelet). By assessing the prediction performance of all single features, the top three predictor features were Max_FFT, MeanDeviation_GLDM, and Kurtosis_Wavelet. Considering the nature of top features in the COVID-19 category, mostly recorded in the frequency domain, It is likely that the change of the variance in the frequency domain is the characteristic feature of the CXR image of COVID-19 cases. In addition, if we averaged the performance of the features in each of the five different groups, the FFT features have better predictive power than the other groups associated with COVID-19. It shows the significance of acquiring such frequency domain features and implies that those features are relevant to the detection of COVID-19 infection in the CXR image.

Third, since identifying optimal and most effective image features is one of the most important and challenging tasks in developing machine learning-based classifiers, we investigated the influences of applying a dimensionality reduction method to select optimal and more correlated features. Interestingly, the results demonstrated that our dimensionality reduction method not only reduces the dimension of feature space but also is able to reorganize the new smaller feature vector with more correlated information and a lower amount of redundancy. Besides, based on the machine learning theory, increasing the ratio of the number of cases per class to the number of features will improve the robustness of the machine learning classifier and reduce the risk of overfitting. Therefore, using this optimal feature selection method, we were able to use a relatively small dataset of 420 cases for the final classifier model, which avoids the large dataset requirement when developing the deep learning-based schemes with the same or even lower accuracy [19].

Despite the encouraging results, we recognize that this study has a few limitations. First, our CXR dataset has a relatively small size. A larger dataset consisting of the cases from different institutions would be useful to further verify the reliability and robustness of our proposed model. Second, in our future work, we will investigate different feature selection and feature reduction methods such as DNE [20], Relief [21], LPP [5], Fast-ICA [22], recursive feature elimination [23], variable ranking techniques [24], or combining them with our feature reduction approach. Third, this study used a neural network-based classifier that can solve complex problems and get adapted well to high dimensional data. However, there may exist needs to explore other effective classifiers such as SVM [25], GLM [26], Random Forest [27].

## Method

### Dataset and Code (GitHub page)

Our Python codes and dataset are available for download on our GitHub page https://github.com/abzargar/COVID-Classifier.git.

This resource is fully open-source, providing users with Python codes used in preparing image datasets, feature extraction, feature evaluation, training the ML model, and evaluation of the trained ML model. We are using a dataset, which is collected from two resources of [28, 29]. Our modified dataset includes 420 2-D X-ray images, in the Posteroanterior (P.A.) chest view, classified by valid tests to three predefined categories of Normal (140 images), pneumonia (140 images), and COVID-19 (140 images). We set all image sizes to 512×512 pixels. Supplementary Figure 2 shows three example images.

### Feature extraction

We used a scheme to compute a total of 252 features in both the spatial and frequency domain. We categorized them into five groups, including Texture [30], Gray-Level Co-Occurrence Matrix (GLCM) [31], Gray Level Difference Method (GLDM) [8], Fast Fourier Transform (FFT) [32], and Wavelet transform [33] as illustrated in Fig. 2. We implemented GLCM and GLDM methods in four different directions, and Wavelet transforms in eight sub-bands. As shown, for each group or each subsection, we computed 14 features by applying the same statistical measures. The 14 features we measured consisted of Mean, Std, Skewness, Kurtosis, Energy, Entropy, Max, Min, Mean Deviation, Median, Range, RMS, Uniformity, MeanGradient, and StdGradient. The feature extraction scheme resulted in 252 features for each X-ray image in total (14 features from Texture, 14 features from FFT, 56 features from GLCM, 56 features from GLDM, and 112 features from Wavelet).

### Evaluation of extracted features’ classification power

Supplementary Figure 3A compares the AUC values among different single features (e.g., Mean, Std_FFT, and Min_Wavelet) for three positive class labels. All features were sorted using the AUC value as an indicator of feature discrimination power. As seen in all three graphs, more than 100 features recorded AUC values higher than 0.6 while features Max_FFT, MeanDeviation_GLDM, and Kurtosis_Wavelet are the top three performers associated with positive class labels of COVID-19, Normal, and Pneumonia with AUC value of 0.87, 0.91, and 0.88, respectively.

Supplementary Figure 3B also shows the performance of five groups of features (e.g., Texture, FFT, and Wavelet) by comparing their average AUC values. As seen, there is no significant difference between them, particularly where the positive label is pneumonia. Given COVID is the target class, the FFT group recorded the best performance, while the best group for the Normal class is GLDM.

### Model training hyperparameters and run-time

For the training process of the designed multi-layer neural network, we chose Adam optimizer to optimize model weights and minimize the categorical cross-entropy loss function. The learning algorithm hyperparameters were set as follows: BatchSize=2, MaxEpochs=100, LearningRate=0.001, DropoutValue=0.2, TrainRatio=0.6, ValRatio=0.2, and TestRatio=0.2. We also used the Early Stopping technique to stop training when the validation score stops improving, aiming to avoid overfitting. The run-time of different parts of our proposed machine learning scheme listed in Table 2, indicates that our model needed a short time of 15.4 seconds to learn, and also predicting one test sample took 2.03 seconds.

**Table 2:** Run-time analysis on the local system with the CPU of Intel Core i7-8750H 2.2 GHz and GPU of RTX2080 Max-Q

|  | Training phase | One single predict phase |  |  |
| --- | --- | --- | --- | --- |
|  |  | Feature Extraction (Fig 1A) | Feature Reduction (Figure 1B) | Classifier (Figure 2) |
| Run-time (Sec) | 15.4 | 1.98 | 0.02 | 0.03 |

## Data Availability

Our dataset and codes are all available on our GitHub page.

https://github.com/abzargar/COVID-Classifier.git

## Acknowledgment

This work was supported by the NIGMS/NIH through a Pathway to Independence Award K99GM126027 (S.A.S.) and start-up package of the University of California, Santa Cruz.

## Author Contributions

A.Z.K and S.A.S designed the project and wrote the manuscript. A.Z.K wrote the classifier and implemented the machine learning code. M.H collected the dataset and wrote the image preprocessing code. This work was supported by the NIGMS/NIH through a Pathway to Independence Award K99GM126027, NIH(NIGMS) (S.A.S.), and start-up package of the University of California, Santa Cruz.

